# Why mothers continue to die in Pakistan: a nested case-control study of predictors of maternal mortality

**DOI:** 10.1101/2023.08.10.23293928

**Authors:** Ahsan Maqbool Ahmad, Iqbal H. Shah, Ali Muhammad Mir, Maqsood Sadiq, Muddassir Altaf Bosan

## Abstract

**Background:** Maternal mortality ratio (MMR) declined in Pakistan from 276 maternal deaths per 100,000 live births in 2006-07 to 186 (confidence interval: 138-234) in 2019. Despite this decline, reasons why mothers continue to die, the inequity in the burden of maternal mortality and its predictors largely remain unknown. We investigate the levels and predictors of maternal mortality in Pakistan.

**Methods:** The in-depth analysis of Pakistan Maternal Mortality Survey (PMMS) 2019 was undertaken using the nested case-control design. We matched 147 maternal deaths that occurred during three years prior to PMMS 2019, defined as “cases”, with 724 women who delivered during the same period, defined as “controls”. We compared socio-demographic background characteristics of cases and controls and performed multivariate regression to investigate the predictors of maternal mortality in Pakistan.

**Results:** Cases and controls were similar on access to antenatal care (ANC) and ANC provider, but differed on age, education, number of pregnancies, type of delivery, tetanus toxoid vaccination during last pregnancy, and ever using a contraceptive method. More of the cases had their last delivery by a skilled birth attendant (83% compared to 63% among controls) and delivered at the government hospital (43% compared to 33% among controls) while home delivery was relatively more common among controls (32% compared to 25% among cases). Odds of maternal death were lowest for women giving birth during age 20-29 (odds ratio – OR: 0.5; 95% CI 0.23-1.07) as compared to women in age 15-19 and those age 30-39 (OR: 1.34; 95% CI 0.61-2.95) and 40-49 (OR: 1.21; 95% CI 0.42-3.45), and among women with secondary or higher education (OR: 0.35; 95% CI 0.17-0.74) compared to women with no education. Surprisingly, women who had their delivery by a skilled birth attendant experienced higher odds of maternal death (OR: 4.07; 95% CI 2.19- 7.57) compared to those who did not. Whereas women having had tetanus injection during last pregnancy (OR: 0.1; 95% CI 0.0-0.96) and ever-used contraceptives (OR: 0.23; 95% CI 0.11-0.39) tend to have reduced odds of maternal death compared to women who did not have the injection and who never used contraceptives, respectively.

**Conclusions:** Women in certain sub-groups confront higher risks of maternal death. Increasing female education, preventing early and late childbearing through contraceptive use, increasing tetanus vaccination during pregnancy, improving providers’ skills, and quality of health care are required to eliminate preventable maternal deaths in Pakistan.

## Introduction

Childbirth is a joyful occasion, yet 287,000 (uncertainty interval – UI: 273,000 to 343,000) women were estimated to have died globally of maternal causes during pregnancy, childbirth or within 42 days of pregnancy termination in 2020 [1]. Despite an overall reduction of 66.9 percent in maternal mortality ratio (MMR) from 417 in 2000 to 138 maternal deaths per 100,000 livebirths in 2020, South Asia accounted for 16.4 percent of all maternal deaths in 2020 globally [1]. Pakistan’s MMR in 2020 was estimated to be higher at 154 compared to other Muslim majority countries of Bangladesh (123), Iran (22) and Egypt (17) and South Asian countries of India (103) and Sri Lanka (29) [1]. Pakistan witnessed an overall decrease in the MMR, from 276 as estimated from the 2006-07 Pakistan Demographic and Health Survey (PDHS) [2] to 186 (CI: 138-234) as reported in the 2019 Pakistan Maternal Mortality Survey (PMMS) [3]. Note that these estimates refer to a period of three years prior to each survey. These figures represent a one-third decline in the number of maternal deaths between 2007 and 2019. However, given that the Sustainable Development Goal (SDG) target for reducing maternal mortality is to lower the MMR to less than 70 maternal deaths per 100,000 live births by 2030 [1], Pakistan still needs to make substantive progress on this indicator and especially to address the high burden of maternal mortality among some population subgroups. Both PDHS and PMMS provide useful information on the level of maternal mortality, nationally, by region and urban-rural place of residence, but little is known about the contextual, health system, social, and biological factors that play an important role in reducing preventable maternal deaths in Pakistan.

Informed by the previous work on studying determinants of maternal death [4,5,6,7,8], we developed a conceptual framework to examine the predictors of maternal mortality using PMMS data. More specifically, we hypothesize that socioeconomic and demographic characteristics and health seeking behavior and comorbidities jointly or individually influence the peril of maternal deaths in Pakistan and disproportionately affect some subgroups of population (Figure 1).

**Figure 1:**
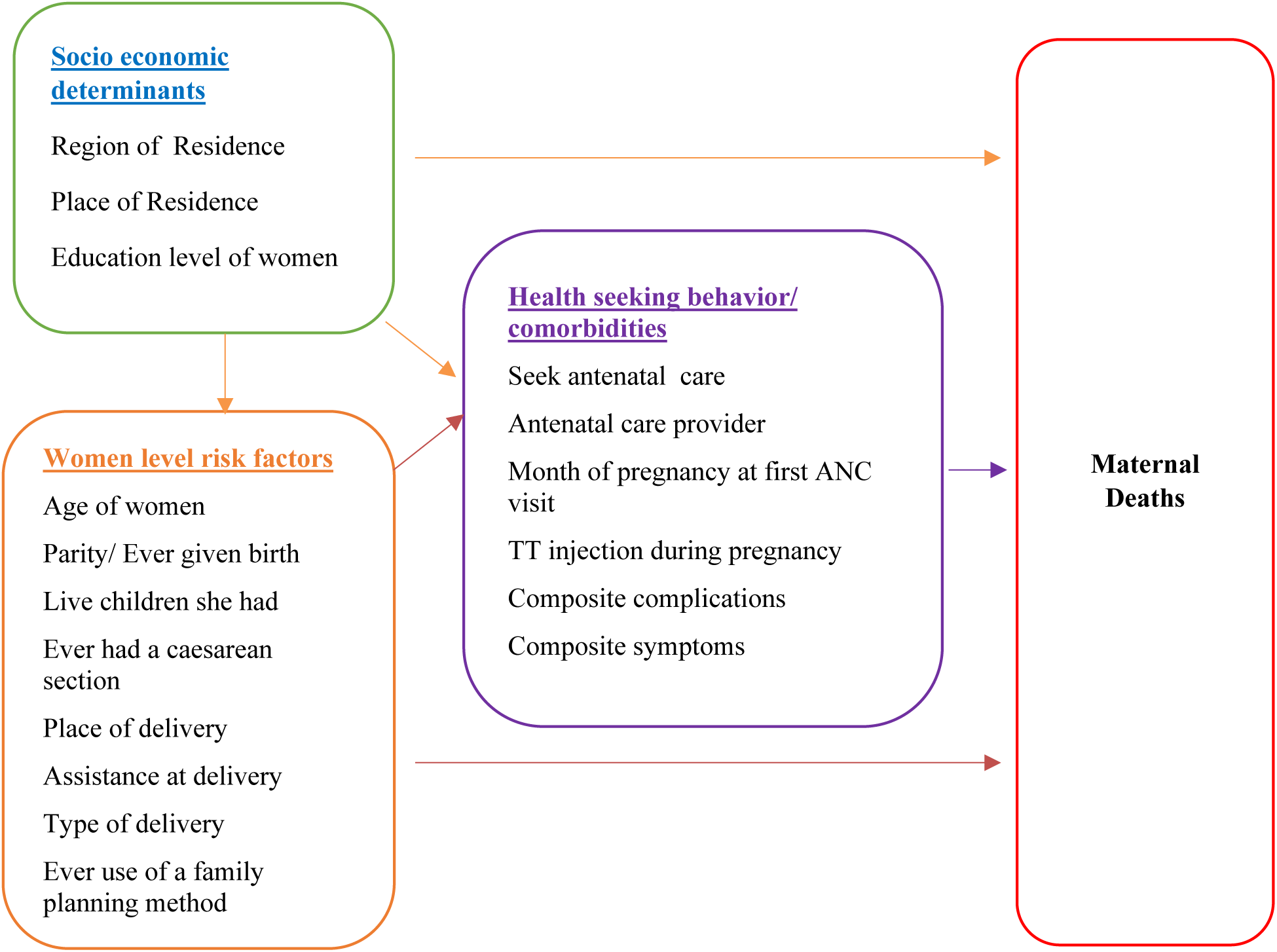
Conceptual framework of predictors of maternal death.

## Materials and Methods

The data were obtained from the PMMS that was the first exclusive survey on maternal mortality in Pakistan. Using a multi-stage cluster design, this nationally representative survey was conducted across four provinces (Balochistan, Khyber Pakhtunkhwa – KP, Punjab and Sindh) of Pakistan as well as in Gilgit Baltistan (GB) and Azad Jammu and Kashmir (AJ&K). We performed a nested case control study in which all direct and indirect maternal deaths (147) identified in the PMMS were regarded as cases, while the controls were randomly selected by matching on cluster from the rest of (6,907) women who reported a live birth during the last three years before the survey. The PMMS Household Questionnaire collected information on all deaths in three years prior to the survey. All female deaths (1,177) were further investigated in detail using the verbal autopsy (VA) questionnaire, which was administered by specially trained interviewers. A total of 1,117 verbal autopsies were reviewed by a panel of expert obstetricians and gynecologists and physicians at the National Committee of Maternal and Neonatal Health (NCMNH). The panel classified maternal deaths into direct, indirect, coincidental, and late maternal deaths, based on the cause of death [9]. International Classification of Diseases 11^th^ Revision (ICD-11) defines maternal death as: the death of a woman while pregnant or within 42 days of termination of pregnancy, irrespective of the duration and site of the pregnancy, from any cause related to or aggravated by the pregnancy or its management but not from unintentional or incidental causes [10]. Direct maternal deaths are those “resulting from obstetric complications of the pregnant state (pregnancy, labour and puerperium), and from interventions, omissions, incorrect treatment, or from a chain of events resulting from any of the above” [10]. Indirect maternal deaths are those “resulting from previous existing disease or disease that developed during pregnancy, and that were not due to direct obstetric causes but were aggravated by the physiologic effects of pregnancy” [10]. Our study analyzes direct and indirect maternal deaths only.

The nested case control design extends the flexibility of focusing specifically on those who experienced mortality (cases) and a set of matched controls per case providing the statistical strength to conduct analysis through which variability due to known and unknown risk factors related to various characteristics and health-seeking behaviours can be controlled in an appropriate manner.

There was a 1:5 ratio between cases and controls. All deaths of women of reproductive age that occurred during the last three years preceding the survey and classified as direct or indirect maternal deaths were included as “cases” in the study (N = 147). On the other hand, all women who were alive and reported a live birth during the same period and matched on the same clusters as cases were classified as “controls”. The controls (N = 724) having the required information about risk factors were included for the analysis (Figure 2). The sample sizes were computed at the 95% confidence level, at a power of 90%, an assumed 20% rate of exposure among controls. Note that the information for cases (deceased women) was provided by the household member(s) most knowledgeable about the deceased woman’s symptoms preceding her death and her background characteristics, whereas information about the controls was provided by women themselves.

**Figure 2:**
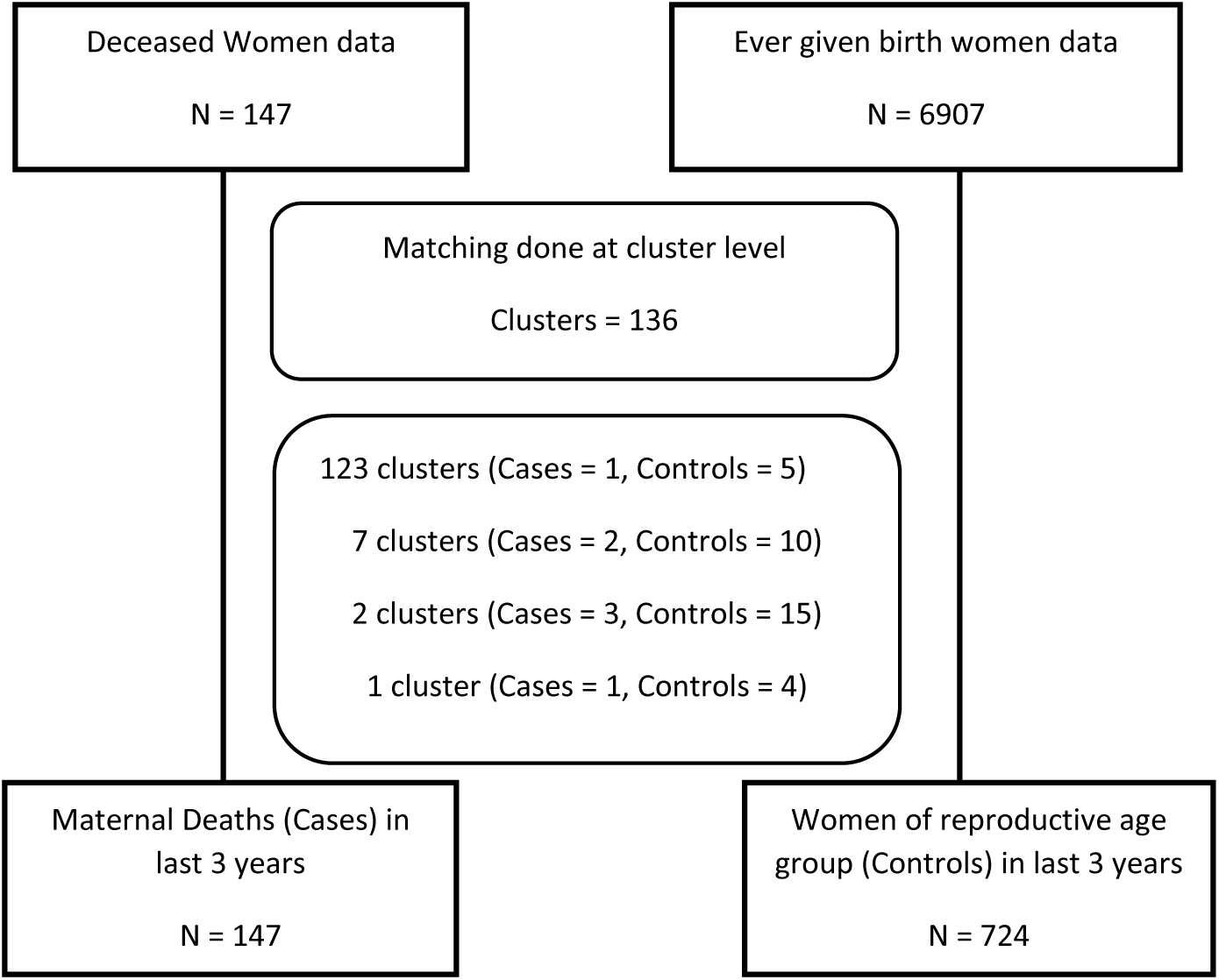
Scheme of cases and controls identification and matching.

## Ethical Statement

We analyzed anonymized data available in the public domain for secondary analysis.

### Statistical Analysis

Analysis was done using Stata version 14 (StatCorp, College Station, Texas). Maternal death was taken as an outcome variable which was dichotomized into whether the death occurred or not (i.e., cases were maternal deaths, and controls were women who were alive and had a live birth). Frequencies and percentages were reported for categorical variables and mean with standard deviation for quantitative variables (for example, composite complications and composite symptoms). Cross tabulations for categorical variables were performed with outcome to identify any sparse data that required merging with other categories.

Across the initial stages of the analyses, three sets of iterations were performed to re-define categories of independent variables. Given the relatively small number of cases in the survey, the number of categories were merged for variables to allow for meaningful comparisons.

To determine the risk factors associated with maternal deaths, these factors were examined separately in univariate analysis. Crude odds ratios (ORs) and matched odds ratios (matched ORs) with 95% confidence interval (CI) were computed by simple logistic regression and conditional logistic regression, respectively. Variables with a p-value less than 0.25 at the univariate level were included in the multivariable analysis and multi-collinearity among the independent variables was checked. Multivariable analysis was done using the same regression techniques to determine risk factors associated with maternal deaths by computing adjusted odds ratios (adjusted ORs) with 95% CIs. Variables having p-value less than 0.05 were kept in the final multivariable model and the Hosmer Lemeshaw goodness of fit test was applied to check goodness of fit of the model using simple logistic regression technique.

We assessed the impact of each of the specified risk factors on maternal mortality by estimating odds ratios (ORs) using a logistic regression model to control for the effects of known biological and socioeconomic variables shown in Figure 1.

## Results

Multi-stage analyses included the phases of descriptive analysis and inferential analyses by applying univariate and multivariate regression techniques. The proportional representation of cases versus controls was similar for the matching variables of region/province of residence and place of residence (i.e. urban vs rural).

Similar proportion of cases and controls who had a live birth, stillbirth, miscarriage or abortion in three years prior to the survey accessed antenatal care (ANC); and saw an obstetrician/gynecologist or a doctor for ANC (Table 1). Cases were, however, distinctly different from controls on important attributes like age group, level of education, number of children alive, type of delivery, tetanus toxoid vaccination during last pregnancy, ever used a contraceptive method. Controls were more numerous in age group 20-29 (54.1%) compared to cases (34.7%), but less numerous in the youngest age group 15-19 (5.9% vs 10.9%) and in older age groups of 30-39 and 40-49. More of controls had middle or less (23.3%) or secondary and higher education (18.8%) compared to cases (17.7% and 12.9%, respectively). More among cases (19.5%) had 7 or more living children than among controls (11.6%). Nearly all controls had tetanus injection (99.9%) compared to cases (86.4%) and more of them ever used an contraceptive method (36.3%) than cases (11.6%) and more of them ever had cesarean section (27.9%) than the controls (14.4%).

**Table 1:**
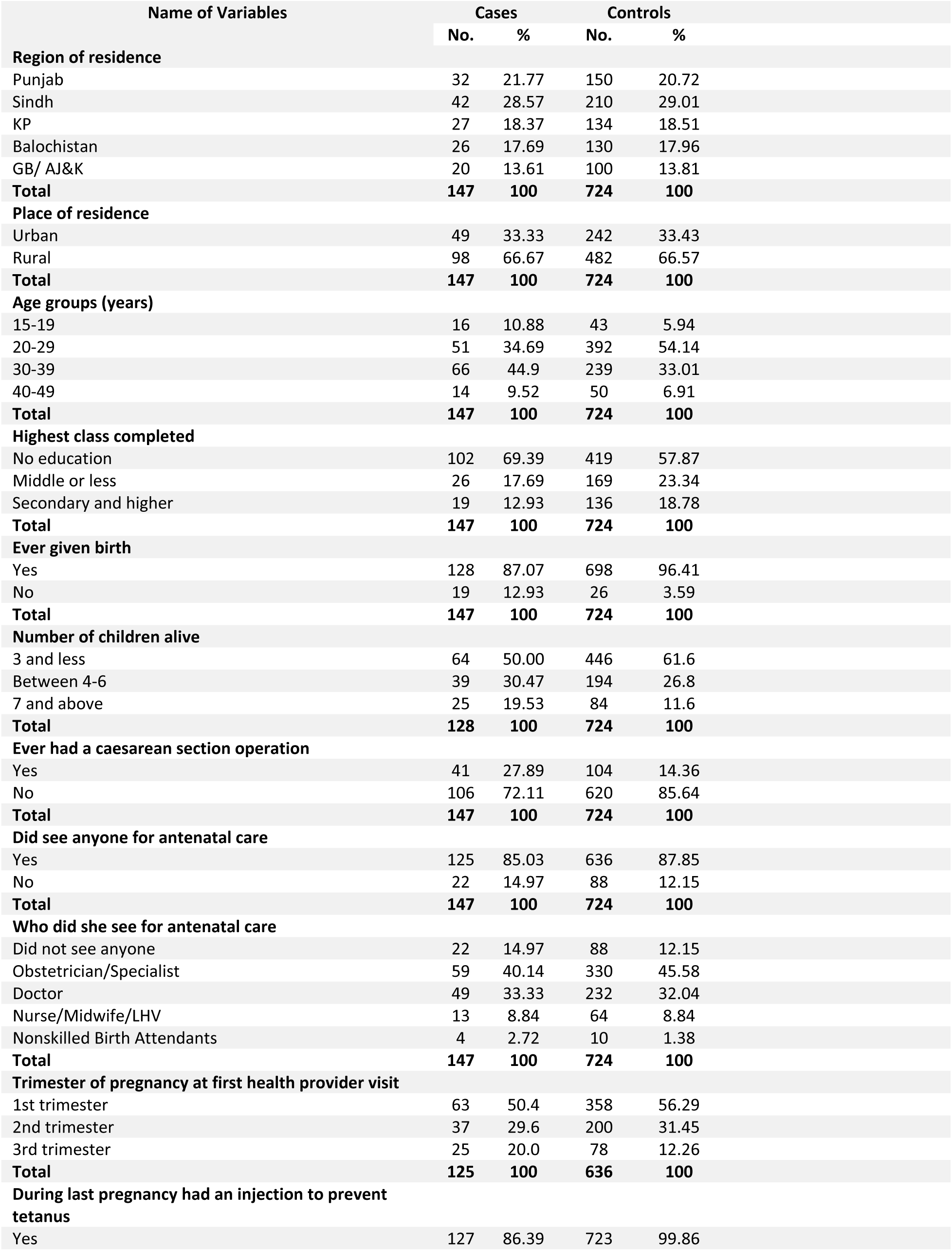
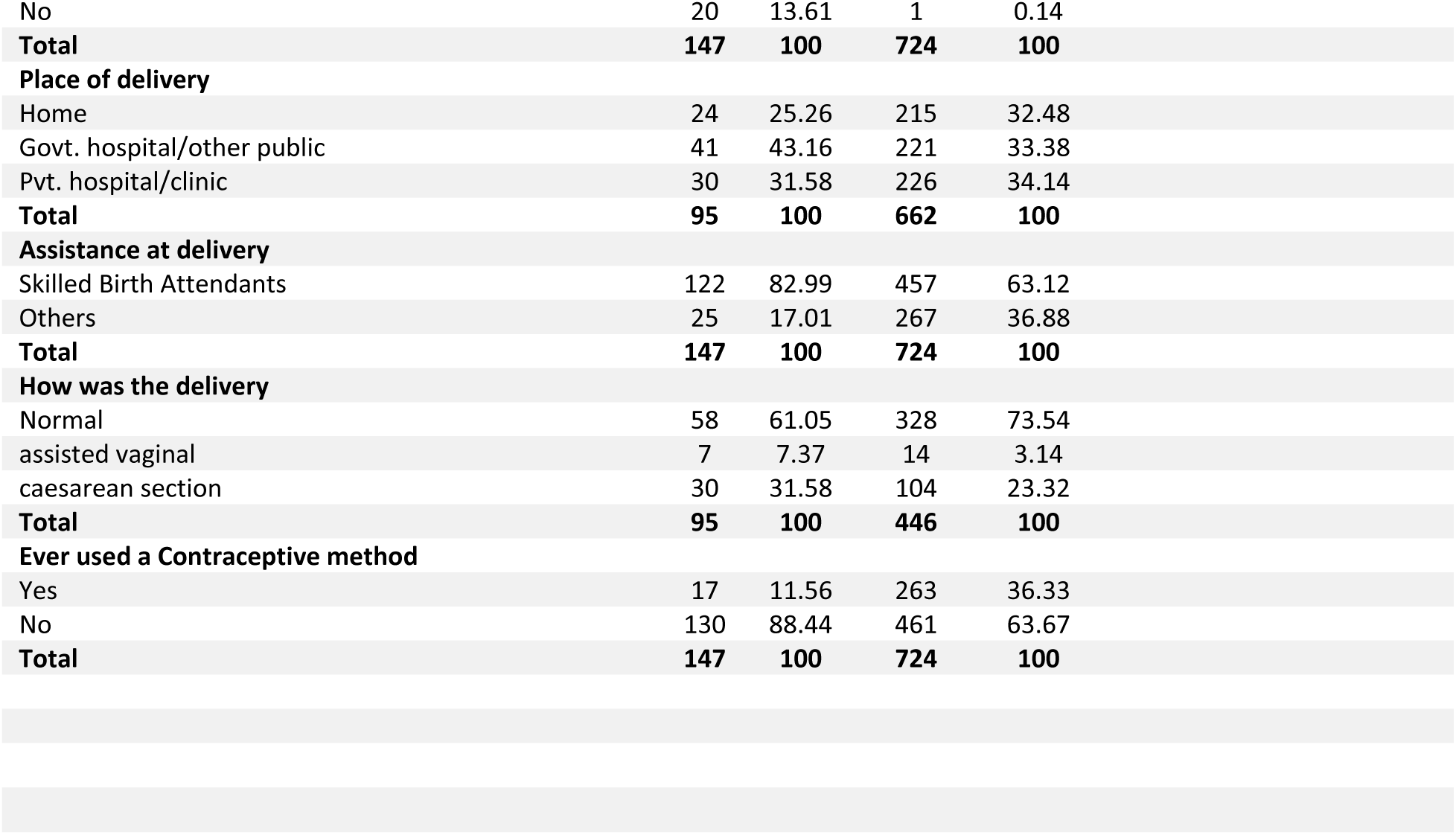
Number and percentage distribution (%) of cases and controls by background variables, Pakistan, 2019.

Surprisingly, more of the cases than controls had their last delivery by a skilled birth attendant (83% vs 63.1%) and delivered at a government hospital (43.2% vs 33.4%) while home delivery was relatively more common among controls (32.5%) compared to cases (25.3%). The average level of composite complications index (including high blood pressure, diabetes, anemia and jaundice during last pregnancy) was similar - 0.62 (standard deviation 0.77) for cases as compared to 0.63 (standard deviation 0.77) for controls. However, cases experienced a higher level (2.35 with standard deviation 1.86) of composite symptoms (fever, fits, vaginal bleeding, jaundice, abdominal pain, breathing difficulty, paleness/anemia, swelling feet or ankles and swelling face during last pregnancy/illness) compared to controls (1.85 with standard deviation 1.62).

The multivariate analysis controlled for the significant confounding effects of age, education, ever given birth, ever had a cesarean section, had injection in the last pregnancy to prevent tetanus, whether received delivery assistance from a skilled attendant, and ever used a contraceptive method while looking at the conditional odds of each one for maternal death (Table 2). The odds ratios after controlling for factors included in the model, were higher for women giving birth at younger age (15-19) or at older ages (30-39 and 40-49). Age group 20-29 had the least risk of 0.5 adjusted odds ratio (CI: 0.23-1.07) compared to women in ages 15-19. Women with education had lower odds than those with no education with 0.35 odds ratio (CI: 0.17-0.74) of maternal death for women with secondary education and 0.51 (CI: 0.27-0.98) for those with middle or less education compared to 1.0 for women with no education. Having ever had a cesarean section doubles the odds of maternal death while having had the tetanus injection in the last pregnancy drastically reduced the odds for maternal death close to zero as compared to women who were not vaccinated. Also, having had ever used a contraceptive method reduces the odds to 0.21 (CI: 0.11-0.39) compared to 1.0 for never users. Delivery by skilled birth attendant was associated with higher odds of maternal death. Having delivery by a skilled attendant had the matched conditional odds of 4.07 (CI: 2.19-7.57) compared to the reference category of delivery by “others”, including home delivery. Women with prior history of symptoms and those who have caesarean section are more likely to have delivery by a skilled attendant and are thus predispose to the risk of death, especially if the quality of care is poor or the attendant lacks the required skills.

**Table 2:** Multivariate conditional logistic and unconditional logistic regression of predictors of maternal mortality, Pakistan, 2019.

| Name of Variables | Conditional logistic regression<br>(N=871) |  |  | Unconditional logistic regression<br>(N=871) |  |  | Goodness of fit |
| --- | --- | --- | --- | --- | --- | --- | --- |
|  | Matched<br>Adjusted<br>OR's | 95% CI's | *p*<br>value | Adjusted<br>OR's | 95 % CI's | *p* value |  |
| Age groups (years) |  |  |  |  |  |  |  |
| 15-19 | 1 | - | - | 1 | - | - | N= 871<br>Hosmer and<br>lemeshow Goodness<br>of fit – test statistics<br>(chi²) = 62.39<br>p value = 0.9380<br>(model is good fit) |
| 20-29 | 0.50 | (0.23 – 1.07) | 0.075 | 0.47 | (0.21 - 0.93) | 0.031 |  |
| 30-39 | 1.34 | (0.61 – 2.95) | 0.467 | 1.24 | (0.59 – 2.61) | 0.564 |  |
| 40-49 | 1.21 | (0.42 – 3.45) | 0.726 | 1.06 | (0.40 – 2.83) | 0.912 |  |
| Highest class completed |  |  |  |  |  |  |  |
| No education | 1 | - | - | 1 | - | - |  |
| Middle or less | 0.51 | (0.27 – 0.98) | 0.042 | 0.57 | (0.33 - 0.99) | 0.046 |  |
| Secondary and higher | 0.35 | (0.17 – 0.74) | 0.006 | 0.49 | (0.27 – 0.90) | 0.021 |  |
| Ever given birth |  |  |  |  |  |  |  |
| Yes | 0.19 | (0.09 – 0.39) | 0.000 | 0.21 | (0.10 – 0.45) | 0.000 |  |
| No | 1 | - | - | 1 | - | - |  |
| Ever had a caesarean section operation |  |  |  |  |  |  |  |
| Yes | 2.05 | (1.18 - 3.55) | 0.011 | 2.22 | (1.35 - 3.66) | 0.002 |  |
| No | 1 | - | - | 1 | - | - |  |
| During last pregnancy had an injection to prevent tetanus |  |  |  |  |  |  |  |
| Yes | 0.007 | (0.001 – 0.06) | 0.000 | 0.005 | (0.001 – 0.046) | 0.000 |  |
| No | 1 | - | - | 1 | - | - |  |
| Assistance at delivery |  |  |  |  |  |  |  |
| Skilled Birth Attendants | 4.07 | (2.19 – 7.57) | 0.000 | 4.13 | (2.32 – 7.35) | 0.000 |  |
| Others | 1 | - | - | 1 | - | - |  |
| Ever Used a Contraceptive method |  |  |  |  |  |  |  |
| Yes | 0.21 | (0.11 - 0.39) | 0.000 | 0.22 | (0.12 - 0.39) | 0.000 |  |
| No | 1 | - | - | 1 | - | - |  |
| *\*Matching variables in relation to urban – rural status and province of residence were retained in the model as matching variables* |  |  |  |  |  |  |  |

*\*Matching variables in relation to urban – rural status and province of residence were retained in the model as matching variables*

## Discussion

Despite the decline in the levels of maternal mortality, substantial inequalities continue to persist by region, urban-rural place of residence and by subgroups of population in Pakistan [9]. The greatest decline from 2006-07 to 2019 was observed in Balochistan province which nevertheless continues to exhibit the highest MMR of 298 (CI: 130-466) per 100,000 live births in 2019 compared to any other region [3]. Compared to 2006-07, progress was also noted in 2019 for an increase in literacy rate and higher educational attainment, especially in rural areas [9]. Higher order (6 or more) births declined from 22% to 15%; four or more ANC visits nearly doubled from 28% to 52% as well as the visit to obstetrician/gynecologist and doctor for ANC from 33% to 60% [2, 3]. Not having even one ANC visit during the last pregnancy declined from 35% to 8%. The coverage of mobile phones in rural areas also doubled from 46% in 2006-07 to 93% in 2019 [2, 3].

This progress has been, however, slow and uneven. Women in rural areas, with no education and those living in Balochistan province continue to suffer excessive risk of maternal death than their counterparts in urban areas, living in other regions of Pakistan or educated, especially those attaining higher levels of education. In addition, little progress has been made in rural infrastructure in terms of improved access to the nearest functioning basic health unit (BHU), rural health centre (RHC), secondary/tertiary hospital or the availability of motorized public transport [9].

The findings that women in the age group of 20-29, with education, had ever used a contraceptive method, or had tetanus injection during the last pregnancy had lower odds of maternal death are expected and consistent with findings from other studies both in Pakistan and elsewhere, The finding that women whose last delivery was by a skilled birth attendant had higher odds of maternal death is counterintuitive, though consistent in both PDHS 2006-07[8] and our analysis of PMMS data. There are three explanations for this unexpected result derived from the in-depth analysis of the verbal autopsies of the deceased women identified in the PMMS [9]. First, women after having sought care from various types of care providers for symptoms/complications of pregnancy reached the skilled attendant with a severe complication/symptom(s) in a critical condition in many cases after shuffling between two to three facilities before reaching the final referral facility. The in-depth analysis further showed that majority of maternal deaths occurred at health facilities following delays in deciding to seek professional care and in reaching an appropriate health facility for care [9]. In a social-cultural context with little or no birth planning and accessing care when situation is worsened, this selectivity in terms of women with a complicated pregnancy or delivery reaching late a skilled attendant, manifested in a higher risk of death. Also, women with prior history of symptoms or those requiring cesarean section are more likely to approach skilled attendant for delivery. Such a pattern is noted where effective referral system is not operational. A study of rural areas of Balochistan and North West Frontier Province (now renamed as Khyber Pakhtunkhwa) showed a similar pattern of high risk of maternal mortality for women who had delivered by a skilled attendant compared to those by a family member or traditional birth attendant [11]. A similar pattern was also observed in eight urban squatter settlements of Karachi [12].

Second, it was found that skilled attendants were not adequately trained and provided a sub-optimal quality of care. The in-depth analysis shows that 36% of all maternal deaths, direct or indirect, and 91% of direct maternal deaths were due to surgical or medical misadventures [9]. Indifference by unmotivated staff, poor skills of health care providers, and lack of medicine and equipment together contributed to heavy death toll of women reaching the facility in reasonable condition [9]. Third, the finding of higher maternal mortality among women with births attended by a skilled provider is consistently reported in studies from Pakistan and the qualitative analysis of verbal autopsies indicates that women with complicated pregnancy or delivery to start with were more likely to seek care from a skilled attendant. The pathways to skilled delivery is, therefore, shaped by the prior history of symptoms exacerbating pregnancy complications leading to institutional care and delivery by a skilled attendant. In addition, the poor quality of service, including poor skills of providers contribute to higher maternal mortality for women with delivery by a skilled attendant. This noteworthy finding indicates that the emphasis on institutional deliveries and delivery by a skilled birth attendant to reduce maternal mortality are insufficient to reduce maternal mortality where norms for pregnancy care are lacking, but a necessary concomitant of complicated pregnancy. Further, quality of care and strengthening providers’ skills must be ensured to provide the care women need on reaching the facility.

Estimating maternal mortality and its risk factors require an exceptionally large sample size that is often not feasible. To keep the sample size within manageable limits, a three-year recall of births and deaths was used in PDHS 2006-07 and PMMS. This approach has problems in that the recall of deaths may have declined considerably during the second and third years before the survey, presumably due to recall errors, misreporting of dates, and/or dissolution or change in the composition of households. It is, therefore, possible that recall errors for the second or third year before the survey underreported maternal deaths. Also, information on the causes and circumstances of deaths ascertained through the verbal autopsy may be less reliable for deaths that occurred in the second or third years of recall.

Another potential limitation is the intentional or unintentional misreporting. Given the cultural sensitivity, induced abortions are notoriously underreported or misreported, especially in the restrictive legal contexts such as Pakistan. It is, therefore, possible that some of the induced abortions and related deaths were under- reported.

## Conclusions

Despite the progress made in reducing maternal mortality, approximately 1 in 143 women in Pakistan will die during her lifetime due to complications during pregnancy, childbirth/abortion, or during the 42 days following pregnancy termination. WHO estimated that 98,000 maternal deaths occurred in Pakistan in 2020 [1]. High levels of maternal mortality, inequity in the burden of maternal mortality and the finding that women having delivery by a skilled attendant suffer higher risk of death indicate a number of policy and programmatic implications. First, health system needs to improve the quality of obstetric care provided especially within the public sector facilities. The government must properly equip facilities and institute proper accountability mechanisms so that all deaths are audited and accounted for. Second, pre-marital counselling should be launched to discourage childbearing during adolescence. Both young (<20 years) and older women of ages over 30 should be given priority attention during ANC, delivery and the postnatal period. Third, family planning should be promoted for maternal and child survival through birth spacing and prevention of unintended and high-risk pregnancies. According to the PDHS 2017-18, nearly 18% of women became pregnant six to 17 months after a live birth and 37% within 24 months. PDHS data show that the highest proportion of closely spaced pregnancies occur in the adolescent age group of 15-19 years. Compared to older women, this group also has the highest unmet need for birth spacing—nearly one out of six women in this age bracket wants to space their pregnancies but is unable to do so.

Unmet need for family planning (FP) stems from the inability of women to access services. The Lady Health Worker (LHW) program, launched in 1994, had the mandate to provide doorstep FP services to rural women, who have higher unmet need and more unintended pregnancies than urban women. This highly acclaimed program is currently plagued with many issues, the foremost of which is a perennial shortage of contraceptive supplies. Within public sector health facilities, service providers do not consider offering family planning services and counselling as their responsibility. This needs to be changed through some bold decisions by the government. A major step should be mandatory provision of FP services through the public and private health networks. This has several advantages, including avoiding the hesitancy some women and men have in visiting the socially stigmatized Family Planning centers run by the Population Welfare Department. General health facility visits offer several opportunities for discussing family planning, such as during antenatal care visits, immediately after delivery, during postnatal checkups, and child immunization visits. On these occasions, couples can be encouraged to discuss their family planning needs and individually focused options can be suggested to help meet their needs. This service delivery approach would also allow men to discuss family planning with male health care providers.

The Population Council estimates that even without increasing the current coverage of skilled birth attendance, by simply meeting the 17 % unmet need for family planning and thus raising current contraceptive use from 34% to 51%, Pakistan could lower maternal mortality by around 30% and save every year nearly 4,000 maternal lives.

Third, tetanus toxoid injection during pregnancy should be universal because of its protective effect. Pakistan must strengthen its immunization program and ensure that the injection is made available at both public and private sector facilities. Fourth, a functional and efficient referral system needs to be in place that prevents women shuffling between facilities and instead reach an appropriate facility equipped to provide comprehensive obstetric care. For ensuring the functional integrity of the referral system, an ambulance system must be put in place that connects the lower to the higher referral facilities. Interventions that make optimal use of the high coverage of mobile phone with home visits by Lady Health Workers and Community Midwives are needed for the continuum of care for maternal, neonatal and child health. The emphasis on institutional delivery and by a skilled birth attendant must be accompanied with strengthening quality of care and providers’ skills, especially in public sector hospitals and facilities that are accessed by poor women and those with little or no education. Concerted efforts and investments are urgently needed to meet the public health and human rights imperative of saving maternal lives. This will also enable Pakistan to meet the Sustainable Development Goal 3 target 3.1 to reduce MMR to less than 70 per 100,000 live births by 2030.

## Data Availability

The data underlying the results presented in the study are available from https://dhsprogram.com/data/available-datasets.cfm

## Author Contributions

AMA, AMM, and IHS conceived the idea for the study developed the initial plans, and drafted the manuscript. MS created data files and supported analysis of data and MAB conducted data analysis. All authors have contributed to the final version of the manuscript.

## Acknowledgements

We thank National Institute of Population Studies for providing the PMMS data and members of the Technical Advisory Committee for helpful comments and suggestions.

## References

1. World Health Organization (WHO). Trends in maternal mortality 2000 to 2020: Estimates by WHO, UNICEF, UNFPA, World Bank Group and the UNDESA/Population Division. Geneva: World Health Organization, 2023.

2. National Institute of Population Studies (NIPS) [Pakistan], and Macro International Inc. Pakistan Demographic and Health Survey 2006-07. Islamabad, Pakistan: National Institute of Population Studies and Macro International Inc, 2008.

3. 3. National Institute of Population Studies (NIPS) [Pakistan] and ICF. Pakistan Maternal Mortality Survey 2019. Islamabad, Pakistan, and Rockville, Maryland, USA: NIPS and ICF, 2020.

4. McCarthy J and Maine Deborah. 1992. A framework for analyzing the determinants of maternal mortality. Studies in Family Planning 1992; 23(1): 23-33.

5. Filippi Veronique, Chou D, Barreix M, et al. on behalf of the WHO Maternal Morbidity Working Group (MMWG). A new conceptual framework for maternal morbidity. Int J Gynecol Obstet 2018; 141 (Suppl. 1): 4–9.

6. Cleland J, Conde-Agudelo A, Peterson H, et al. Contraception and health. Lancet 2012; 380 (9837): 149–156.

7. Hanif M, Khalid S, Rasul A, et al. Khalid Mahmood. Maternal Mortality in Rural Areas of Pakistan: Challenges and Prospects. Chapter in IntechOpen.

8. Midhet F, Durrani S, and Jaffery S. Maternal Mortality. Women and Children’s Health: an in-depth Analysis of 2006-07 Pakistan Demographic and Health Survey Data.

9. The Population Council. Explaining maternal mortality: an in-depth analysis of Pakistan Maternal Mortality Survey 2019. 2022, Islamabad: The Population Council.

10. 10. World Health Organization. International Classification of Diseases 11^th^ Revision (ICD-11). https://icd.who.int.

11. Midhet F, Becker S and Berendes HW. Contextual determinants of maternal mortality in rural Pakistan. Social Science and Medicine 1988, 46 (12): 1587–1598.

12. Fikree FF, Gray RH, Berendes HW and Karim MS. A community-based nested case-control study of maternal mortality. Int J Gynecol Obstet 1992, 47(3): 247–255.

